# Microstructure predicts cognition following deep brain stimulation in Parkinson’s disease

**DOI:** 10.64898/2026.11.19.26368410

**Authors:** Lijiao Chen, Laura Witt, Alexander Calvano, Miriam H. A. Bopp, Christopher Nimsky, Haidar S. Dafsari, Lars Timmermann, David J. Pedrosa, Marcus Belke, Philipp A. Loehrer

## Abstract

Deep brain stimulation of the subthalamic nucleus (STN-DBS) effectively treats motor and non-motor symptoms in advanced Parkinson’s disease (PD). However, 15-20 % of patients experience postoperative cognitive decline and identifying preoperative markers remains an unmet clinical need. In this prospective open-label study, we employed NODDI-DTI to investigate whether cerebral microstructure predicts cognitive outcomes following STN-DBS. Forty PD patients underwent diffusion MRI and neuropsychological assessments at preoperative baseline and 6-month follow-up. A whole-brain voxel-wise analysis related microstructural metrics to postoperative cognitive change. Compromised microstructure within three large-scale networks – the frontoparietal/attentional, default mode/limbic, and ventral associative/visual networks – was associated with greater postoperative cognitive decline. These findings suggest that both disease-related network degeneration and stimulation acting on compromised circuits may contribute to postoperative cognitive decline. In conclusion, preoperative evaluation of microstructural integrity may help identify patients at risk for postoperative cognitive decline, supporting individualised counselling and targeted behavioural monitoring after STN-DBS.

## Introduction

Subthalamic nucleus deep brain stimulation (STN-DBS) is an effective treatment for both motor and non-motor symptoms in Parkinson’s disease (PD) and improves patients’ quality of life.^1^ ^2^ ^3^Cognitive decline, however, affects 15-20 % of patients undergoing STN-DBS and remains one of the most significant factors limiting the beneficial effects on quality of life.^4^ ^5^ While negative postoperative trajectories have been observed for executive function, verbal learning, and memory, there are no validated preoperative predictors that can identify patients at risk, representing an unmet clinical need.^6^

From a neuropathological standpoint, Lewy-related pathology in cortical areas and dopamine deficiency in frontostriatal pathways are central to the development of cognitive impairment in PD.^7^ ^8^ At the functional level, large-scale network disruption – including the frontoparietal, default mode, and posterior cortical networks – is involved in the emergence of cognitive decline rather than isolated regional pathology.^9^ ^10^

Despite this mechanistic understanding, the attribution of postoperative cognitive change to DBS remains contested. Functional connectivity studies have shown that stronger coupling between DBS sites and the anterior cingulate, caudate, hippocampus, and cerebellar cognitive regions predicts DBS-induced cognitive decline.^5^ Yet randomised controlled trials and observational studies have largely failed to detect substantial changes in global cognitive performance, suggesting that postoperative trajectories may reflect preexisting structural and in particular microstructural vulnerability rather than a direct stimulation effect.^11^ ^12^ Resolving this question requires identifying preoperative markers of individual predisposition to postoperative cognitive change.

To predict treatment response, diffusion tensor imaging (DTI) has recently received increasing attention.^13^ ^14^ In particular, quantitative MRI techniques investigating cerebral microstructure in vivo have been employed to track disease progression and postoperative outcomes.^13^ ^15^ Here, neurite orientation dispersion and density imaging (NODDI) is an advanced diffusion MRI technique that characterises white and grey matter microstructure with greater biological specificity than conventional DTI.^16^ It yields two complementary metrics: the neurite density index (NDI), reflecting axonal and dendritic density, and the orientation dispersion index (ODI), capturing the geometric complexity of neurite organisation.^16^ We have previously demonstrated that preoperative microstructure predicts non-motor and motor outcomes following STN-DBS in PD.^13^ ^15^ ^17^ Building on this work, the present study tests whether preoperative microstructure is associated with cognitive change following STN-DBS, with the goal of informing preoperative patient counselling and targeted behavioural monitoring strategies.

## Results

### Clinical outcomes

At the 6-month follow-up, the following outcomes improved (see Table 1): Parkinson’s Disease Questionnaire-8 summary index (PDQ-8 SI; p = .002, Cohen’s d: .53), levodopa equivalent daily dose (LEDD; p < .001, Cohen’s d: 1.0), LEDD of dopamine agonists (LEDD-DA; p < .001, Cohen’s d: .72), Scales for Outcomes in PD-Motor Function (SCOPA-Motor) total score (p < .001, Cohen’s d: .98) as well as its subscores SCOPA motor evaluation (p = .003, Cohen’s d: .64), activities of daily living (p < .001, Cohen’s d: .69), and motor complications (p < .001, Cohen’s d: .73). No group level changes in MoCA were observed (p = .91, Cohen’s d: -.04).

**Table 1:** Clinical characteristics between baseline and 6-month follow-up.

|  | Baseline |  |  | 6-MFU |  |  | Baseline vs. 6-MFU |  |
|---|---|---|---|---|---|---|---|---|
|  | *n* | *M* | *SD* | *n* | *M* | *SD* | *p* | *Cohen’d* |
| MoCA total score | 40 | 26.5 | 3.1 | 40 | 26.62 | 2.74 | .92 | -0.04 |
| SCOPA total score | 38 | 19.66 | 6.59 | 39 | 13.23 | 6.51 | **<.001** | .98 |
| Motor evaluation | 38 | 9.16 | 4.23 | 39 | 6.64 | 3.58 | **.004** | .64 |
| Activities of daily living | 38 | 6.68 | 2.83 | 40 | 4.55 | 3.36 | **<.001** | .69 |
| Motor complications | 38 | 3.79 | 2.36 | 40 | 2.02 | 2.49 | **<.001** | .73 |
| PDQ-8 SI | 40 | 31.56 | 15.52 | 40 | 23.59 | 14.28 | **.002** | .53 |
| LEDD [mg] | 40 | 918.05 | 405.29 | 40 | 569.94 | 282.4 | **<.001** | 1 |
| LEDD DA [mg] | 40 | 257.42 | 131.42 | 40 | 164.33 | 128.34 | **<.001** | .72 |

**Abbreviations:** 6-MFU = 6-month follow-up; MoCA = Montreal Cognitive Assessment; SCOPA = Scales for Outcomes in Parkinson’s Disease; PDQ-8 SI = 8-item Parkinson’s Disease Questionnaire summary index; LEDD = Levodopa equivalent daily dose; LEDD-DA = LEDD of Dopamine Agonists

### Fractional Anisotropy and postoperative cognitive outcomes

Whole brain voxel-wise analysis yielded three significant fractional anisotropy (FA) clusters that were negatively associated with postoperative change in MoCA (cf. Supplementary Table 1, Supplementary Figure 1): left superior longitudinal fasciculus (SLF; N1, N3 p < .001-.033), right superior frontal gyrus (SFG) and middle frontal gyrus (MFG; N2, p = .021).

### Orientation Dispersion Index and postoperative cognitive outcomes

Whole brain voxel-wise analysis revealed four ODI clusters that were positively associated with changes in postoperative MoCA (cf. Supplementary Table 2, Figure 1): left SLF and corticospinal tract (CST; P1, p < .001), right CST, anterior thalamic radiation (AThR) and SLF (P2, p < .001), and right CST (P4, p = .029), right lingual gyrus, cerebellar V, and cerebellar VI (P3, p = .011).

**Figure 1.**
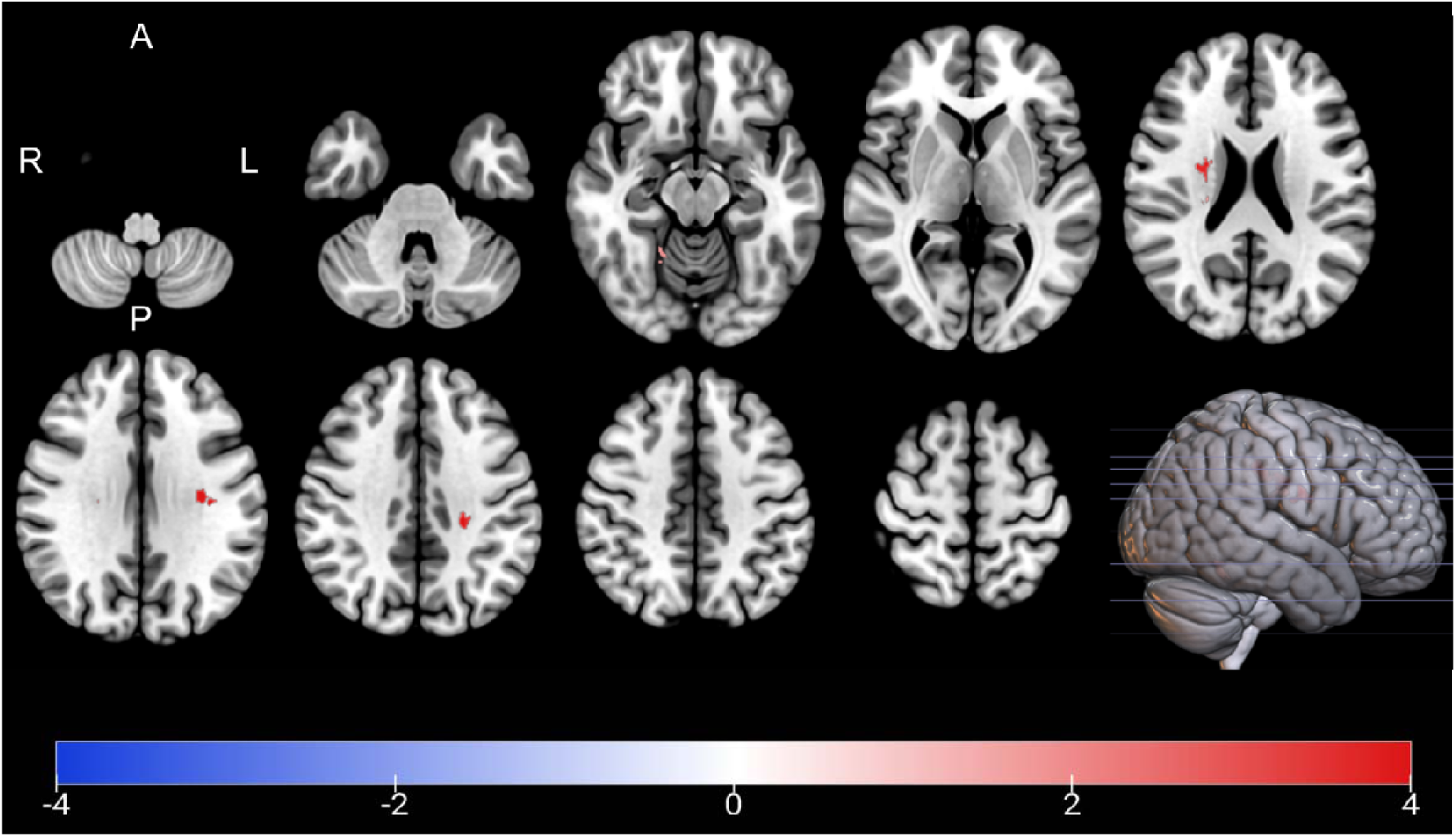
Association between ODI and postoperative change in MoCA. Clusters with a positive association between PD patients’ ODI-values and postoperative change in MoCA as revealed by the whole brain analysis. Clusterwise p-values were corrected for multiple comparisons using a permutation-based approach and shown as the negative decadic logarithm of the p-value.

### Neurite Density Index and postoperative cognitive outcomes

Whole brain voxel-wise analysis revealed seven significant NDI clusters that were negatively associated with change in postoperative MoCA (cf. Supplementary Table 3, Figure 2): left and right lateral occipital cortex (OC; N1, N2, N3, p ≤ .001), left and right inferior fronto-occipital fasciculus (IFOF; N2, p ≤ .001), left and right inferior longitudinal fasciculus (ILF; N2, N3, p ≤ .001), bilateral cingulate gyrus (CG; N4, N6, N7, p = .001-.026), bilateral precuneous (N6, N7, p = .014-.026), and right SLF and CST (N5, p = .002).

**Figure 2.**
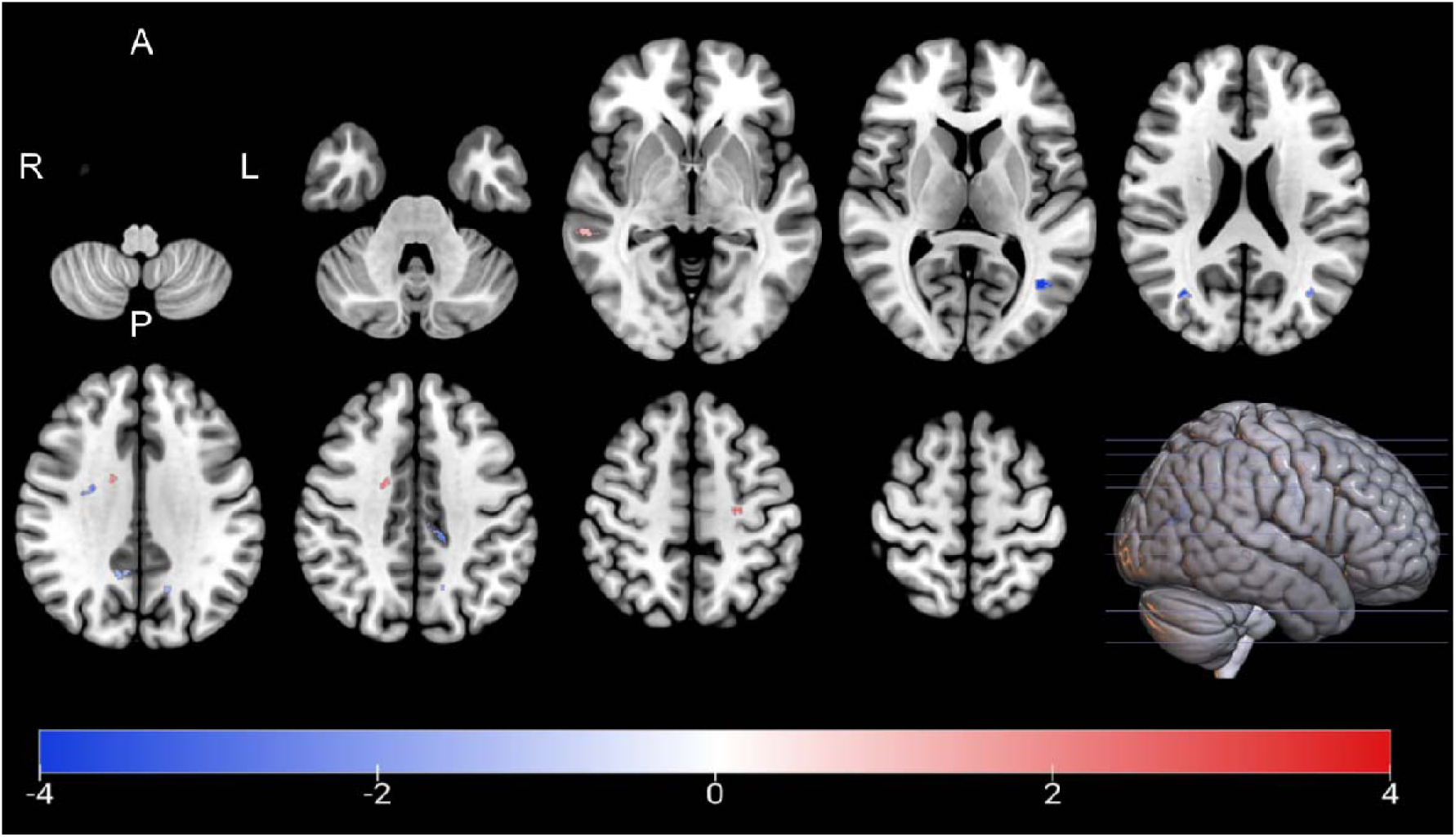
Association between NDI and postoperative change in MoCA. Clusters with a positive and negative association between PD patients’ NDI-values and postoperative change in MoCA as revealed by the whole brain analysis. Clusterwise p-values were corrected for multiple comparisons using a permutation-based approach and shown as the negative decadic logarithm of the p-value.

### Microstructure and postoperative cognitive outcomes adjusted for stimulation overlap

In a supplementary analysis, we repeated the GLM analysis with the overlap between the volume of tissue activated (VTA) and the STN added as a covariate. This tested whether the association between microstructural metrics and postoperative cognitive change depended on the extent of STN stimulation. For conciseness, the results are shown in Supplementary Figures 3-5. Some smaller clusters no longer survived correction for multiple comparisons after this adjustment. The main findings for cognition, however, remained consistent with the primary analysis.

## Discussion

In the present study, we employed NODDI-DTI to investigate whether microstructure is associated with cognitive outcomes following STN-DBS in patients with Parkinson’s disease. Our findings demonstrate that compromised microstructure within the frontoparietal/attentional network, the default mode/limbic network, and the ventral associative/visual network is associated with greater postoperative cognitive decline. These results support two main conclusions. First, they point to two complementary mechanisms influencing postoperative cognitive outcomes. Microstructural impairment in networks typically modulated by DBS suggests that stimulation may contribute to cognitive deterioration when it acts on already vulnerable circuits. By contrast, compromised microstructure in networks not classically targeted by DBS suggests that disease-related degeneration of distinct networks may also contribute to postoperative cognitive decline. Second, preoperative evaluation of microstructural integrity may support clinical decision-making by identifying patients at risk for postoperative cognitive decline, thereby supporting patient counselling and stratification into targeted behavioural monitoring strategies.

In the present study, compromised microstructure as indexed by higher ODI in the frontoparietal/attentional network (SLF, AThR) as well as the motor pathway (CST) was associated with stronger postoperative cognitive decline. In white matter, lower ODI indicates more parallel and coherent fibre orientations, while higher ODI is related to increased fanning of WM tracts, indicating disorganised and degenerated axons.^16^ In PD, the accumulation of alpha-synuclein within axonal compartments is associated with axonal degeneration^18^ and increasing evidence shows that microstructural alterations in white matter are related to neurocognitive and psychiatric disorders in PD.^19^ The identified cluster within the superior subdivision of the left SLF (cluster P1, cf. Supplementary Table 2) corresponds to SLF I which is implicated in spatial processing and motor planning.^20^ In particular, Li and colleagues (2023) found that higher fibre coherence in bilateral SLF I is related to higher precision of visual working memory.^21^ The CST, another major white matter tract identified in the present study, is the major effector pathway for voluntary motor function.^22^ Interestingly, accumulating evidence suggests that the integrity of CST is relevant for non-motor symptoms and, in particular, non-motor outcomes following STN-DBS.^13^ ^23^ Here, an association between impaired microstructure in right CST and depression was observed in patients,^24^ while we demonstrated in a previous study that microstructure in bilateral CST predicts postoperative non-motor burden.^13^ We further identified an association between postoperative cognitive outcomes and AThR, a key pathway involved in executive function and decision-making.^25^ ^26^ This is in line with our previous results linking lower ODI, i.e. intact microstructure, in AThR with positive postoperative outcomes in the attention and memory domain.^13^ These observations are consistent with the macroscale framework of STN-DBS,^13^ whereby stimulation is thought to exert its therapeutic and adverse effects by modulating large-scale networks whose structural integrity depends on long-range white matter pathways.^27^

Lower NDI was associated with greater postoperative cognitive decline across several clusters that map onto three large-scale networks identified in the present study: the default mode/limbic network (bilateral cingulate gyrus and precuneous), the ventral associative/visual network (left and right lateral occipital cortex, bilateral IFOF, and ILF), and the frontoparietal/attentional network (right SLF). NDI reflects the estimated volume fraction of the intracellular compartment per voxel and is considered a marker of neurite density and integrity.^16^ Consistent with this interpretation, healthy brain tissue exhibits higher NDI than tissue from patients with Alzheimer’s disease,^28^ and among healthy older adults, higher NDI has been associated with better executive function.^29^

In the present study, compromised microstructure in the cingulate gyrus and precuneous was associated with worse postoperative cognitive outcomes. Both structures are core hubs of the default mode network (DMN), which supports higher-order cognitive functions such as memory, conceptual processing, self-referential thought, and social cognition.^30^ The identified clusters extended into IFOF and ILF, which are not canonical DMN nodes themselves, but rather structural pathways supporting DMN connectivity. Previous functional MRI studies have shown that patients with PD exhibit reduced deactivation in the posterior cingulate cortex and precuneus during executive tasks such as card-sorting, accompanied by an abnormal reversal of activation-deactivation patterns within the DMN.^31^ Taken together with the present microstructural findings, these previously reported functional abnormalities may have a structural basis: reduced NDI within DMN hubs and their supporting white matter pathways may reflect microstructural degeneration underlying impaired DMN suppression. This suggests that structural damage may contribute to the network-level dysfunction previously reported in PD.^31^

Furthermore, several affected regions identified in the present study are also implicated in visuospatial and language processing, both of which are assessed by MoCA subtests. Notably, Lewy-related pathology in PD extends beyond the initially affected brainstem and limbic structures to involve posterior cortical regions.^32^ A growing body of evidence indicates that Lewy pathology progressively involves posterior parieto-occipital regions in later disease stages,^33-35^ a pattern which is strongly linked to visuospatial impairment in PD.^33^ ^36^ The lower NDI we observed within the posterior DMN and posterior cortical regions may provide microstructural evidence consistent with this posterior pattern of Lewy pathology progression. Notably, the left and right lateral OC and precuneous appear to be remote network nodes modulated by DBS rather than classic direct stimulation targets, and the IFOF and ILF lack robust and validated evidence as primary DBS-affected tracts.^37-41^ In contrast, the cingulate gyrus has been consistently reported as a site of cortical modulation by STN-DBS.^42^ ^43^ Taken together, these findings suggest that postoperative cognitive outcomes are likely to be driven by both, direct modulation of specific networks whose microstructural integrity is pertinent for DBS-network effects and pre-existing, disease-related degeneration of distinctive networks, irrespective of stimulation.

This dual mechanism helps to reconcile the apparent discrepancy outlined in the introduction between connectivity-based predictions of DBS-induced cognitive decline^5^ and the null effects on global cognition reported in randomised clinical trials.^12^ Group-level analyses may obscure clinically meaningful individual variation that becomes detectable only when preoperative structural vulnerability is explicitly modelled, as in the present approach.

Lower FA was associated with stronger postoperative cognitive decline in clusters within the frontoparietal/attentional network, i.e. in left SLF, right SFG and MFG. Higher FA values represent more directional water diffusion, indicating well-myelinated axon bundles, whereas lower FA values reflect more isotropic diffusion, commonly interpreted as axonal alteration.^44^One of the key findings of the present study is the identification of two distinct subcomponents within left SLF associated with postoperative cognitive change. The first subcomponent (cluster N1, cf. Supplementary Table 1) was located in the dorsal part of the left SLF, which functionally connects the SFG and MFG, forming part of the frontoparietal control network associated with attention, working memory, and executive control (SLF II).^45^ ^46^ The second subcomponent (cluster N3, cf. Supplementary Table 1) was located in the left posterior ventral SLF, which is related to language processing (SLF III).^45^ ^46^ Parlatini et al. suggested that SLF III is functionally linked to a ventral non-spatial/motor network, whereas SLF II serves as an intermediate pathway supporting the full range of frontoparietal functions.^47^ This functional distinction may help explain why both SLF subcomponents identified in our study were associated with cognitive change, despite their different anatomical locations within the tract.

Additionally, the identified SFG and MFG are thought to be nodes of the associative loop of basal ganglia-thalamocortical circuits and are structurally connected to frontoparietal pathways that support executive function.^48^ DBS is known to modulate basal ganglia output within this associative loop^49^ ^50^ and to exert downstream effects on frontoparietal pathways.^51^ We therefore hypothesise that preoperative microstructural integrity of the SLF, SFG, and MFG network reflects the structural capacity of the frontoparietal system to compensate for DBS-induced perturbation of associative loop output. Patients with lower preoperative FA in these tracts may have less structural reserve to buffer this perturbation, rendering them more vulnerable to postoperative cognitive decline.

This research has limitations. First, the DTI acquisition protocol (2.0 × 2.0 × 2.0 mm³; 40 diffusion-encoding directions) represented a compromise between spatial resolution, signal-to-noise ratio, and scan duration. Although consistent with established NODDI protocols,^52^ ^53^ higher-resolution acquisitions may improve characterisation of white matter microstructure. This trade-off was necessary because prolonged MRI acquisitions increase the risk of motion artefacts in people with PD. Second, the assumptions of the NODDI-DTI model may represent an oversimplification and could therefore result in reduced specificity. The underlying biophysiological model has, however, been shown to support a realistic approximation to tissue structure at a clinically feasible acquisition time.^16^ Third, the present study assessed general cognitive function using the MoCA, which does not allow for the differentiation of specific cognitive subdomains. Future studies should incorporate domain-specific neuropsychological assessments and include a non-DBS control group to validate and extend these findings.

In conclusion, we describe a distinct profile of compromised microstructure associated with postoperative cognitive decline, including structures of the frontoparietal/attentional network, the default mode/limbic network, and the ventral associative/visual network. We conclude that two complementary mechanisms shape postoperative cognitive outcomes. First, DBS-mediated modulation of compromised networks may contribute to worsened cognitive outcomes, while disease-related network degeneration results in postoperative cognitive decline independent of DBS network effects. Furthermore, we suggest that diffusion MRI can support preoperative patient counselling by identifying patients at risk for postoperative cognitive decline and enables stratification for targeted monitoring strategies.

## Methods

The study was approved by the ethics committee of Philipps-University Marburg (study-number: 155/17) and conducted in accordance with the Declaration of Helsinki.

### Participants

Forty PD patients (11 female, mean age 58.6 ± 8 years) were enrolled in this prospective, open-label study upon written informed consent. Inclusion criteria are composed of indication for DBS lead surgery because of advanced PD according to modern criteria.^54^ Exclusion criteria include pathological MRI finding, the concurrent presence of neurological or psychiatric disorders, or impaired visual or auditory function.

### Clinical Assessments

Clinical assessments were completed preoperatively and six months following DBS surgery including the following scales:

The Montreal Cognitive Assessment (MoCA) consists of seven domains: visuospatial, naming, memory, attention, language, abstraction, and orientation. The total score ranges from 0 (maximum impairment) to 30 (no impairment). The cutoff for mild cognitive impairment is 26, and the cutoff for dementia is 21.^55^

The PD Questionnaire (PDQ)-8 is a short form of the PDQ-39 and measures eight dimensions of quality of life (QoL) in PD patients.^56^ The score is a well-established measure in PD patients undergoing DBS surgery and is reported as a summary index (SI) with a score range from 0 (no impairment) to 100 (maximum impairment).^57-59^

Scales for Outcomes in PD – Motor Function (SCOPA-Motor) consists 3 dimensions: motor evaluation, activities of daily living, motor complications, with subscale scores ranging from 0 (no impairment) to 42, 21, and 12, respectively.^60^

### Image Processing

Image acquisition parameters are described in the Supplementary Methods. For T1-weighted images, the steps of skull stripping, automated Talairach transformation, cortical and subcortical segmentation, intensity normalisation, grey/white matter tessellation, topology correction, and surface deformation on the basis of intensity gradients were completed using FreeSurfer 7.4.1 (http://surfer.nmr.mgh.harvard.edu).^61^ Processing of the diffusion-weighted imaging data was conducted using FMRIB Software Library (FSL) 6.0.5.2 (https://fsl.fmrib.ox.ac.uk/fsl) including Eddy correction, rotation of the diffusion gradient vectors, and brain extraction.^62^ Afterwards, linear regression was employed to calculate the diffusion tensor for each voxel, from which FA was derived. Furthermore, NDI and ODI were derived using NODDI-DTI, a modification of NODDI.^63^ For voxel-wise analysis, FA maps were processed using Tract-Based Spatial Statistics (TBSS). Individual FA maps were first processed and afterwards nonlinearly registered to standard MNI152 space. A mean FA map and a white matter skeleton were subsequently generated and individual FA values were projected onto the mean FA skeleton using a threshold of FA>0.2. Non-FA diffusion metrics, including NDI and ODI, were subsequently processed using the TBSS non-FA procedure and projected onto the same white matter skeleton.^64^ b0-images were visually inspected to confirm that no changes beyond those in the tissue structure contributed to the observed effects.

Subsequently, an affine matrix was created by registering the first b0-image of each scan to the T1-weighted images using a boundary-based method.^65^ Finally, T1-derived segmentations and brain masks were mapped into diffusion space using the inverse of the affine matrix.

To confirm accurate electrode placement and control for the overlap between the VTA and the STN, electrodes were reconstructed in Lead-DBS (default settings; www.lead-dbs.org). Postoperative CT-scans were first linearly coregistered to the preoperative MRI with Advanced Normalization Tools (ANTs; http://stnava.github.io/ANTs/) and subsequently normalised nonlinearly to standard space (ICBM 2009b NLIN Asym). Electrode trajectories and contacts were then reconstructed using the PaCER algorithm (Supplementary Figure 2). Subsequently, patient-specific VTAs were modelled and the mean percentage overlap between the VTA and bilateral STN calculated.

### Statistical analysis

Statistical analysis of clinical outcomes was performed in R (version 4.4.3). Differences between baseline and follow-up were estimated using paired t-test or Wilcoxon test, as determined by Shapiro-Wilk test for normality.^66^ Effect sizes were estimated based on Cohen’s convention.^67^

For imaging analysis, voxel-wise statistical analysis was performed using a general linear model (GLM) including age, sex, and disease duration as covariates. Only voxels classified as brain tissue across all subjects were included in the analysis. To associate metrics of microstructure and cognitive change, percentage differences between baseline and follow-up values were calculated for MoCA total values using the following formula:^68^

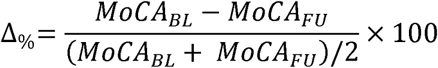

The change in MoCA was associated with FA-, NDI-, and ODI-maps in the voxel-wise GLM. To correct for multiple comparison, a permutation-based approach with 12,000 simulations under the null hypothesis was employed, based on the Analysis of Functional NeuroImages (AFNI) null-z simulator. Clusters were identified based on a voxel-wise significance of p<0.01 and clusterwise p<0.05 was considered significant.

As a supplementary analysis, the GLM was rerun with VTA-STN overlap included as a covariate.

## Data Availability

The data supporting this study’s findings are available on request from the corresponding author (PAL). The data are not publicly available due to privacy or ethical restrictions.

## Code Availability

All tools used to analyse MRI data are based on FreeSurfer Version 7.1 (http://surfer.nmr.mgh.harvard.edu/) and FSL 6.0.5.2 (http://www.fmrib.ox.ac.uk/fsl) packages, which are freely available. Scripts for automation were written in tcshell and parts of the statistics were written in Python using the packages numpy, pandas, seaborn, matplotlib, nibabel and scipy, which are also freely available. Python program code for the analysis of NODDI-DTI is available from https://github.com/dicemt/DTI-NODDI.

## Acknowledgement

The authors thank the participants for their active engagement in this study.

## Contributorship

LC: study concept and design, data acquisition, data analysis, drafting of the manuscript

LW: data analysis, critical revision of the manuscript

AC: data acquisition, critical revision of the manuscript

MBo: data acquisition, surgical intervention, critical revision of the manuscript

CN: data acquisition, surgical intervention, critical revision of the manuscript

HSD: study design, critical revision of the manuscript

LT: study design, critical revision of the manuscript

DJP: data acquisition, drafting of the manuscript

MBe: study concept and design, data acquisition, data analysis, drafting of the manuscript

PAL: study concept and design, data acquisition, data analysis, drafting of the manuscript

