## Supplementary material for "Microstructure predicts cognition following deep brain stimulation in Parkinson’s disease"

**Supplementary Methods**

**MRI Data Acquisition and Processing**

PD patients in the MedON were scanned at baseline on a 3-Tesla Trio scanner (Siemens, Erlangen, Germany) at the Core Unit Brain Imaging of the University of Marburg. The acquisition protocol comprised the following sequences:

1. 3D T1-weighted Magnetization Prepared - RApid Gradient Echo sequence (MPRAGE, field of view (FoV) = 256 mm, matrix 256 x 256, 176 slices, slice thickness 1 mm, voxel dimension 1.0 x 1.0 x 1.0 mm³, repetition time (TR) = 1900 ms, echo time (TE) = 2.26 ms, inversion time (TI) = 900 ms, flip-angle = 9°, bandwidth (BW) = 200 Hz/Pixel, parallel imaging (GRAPPA) with factor 2)
2. diffusion weighted imaging (DWI) (FoV = 256 mm, matrix 128 x 128, slice thickness 2 mm, distance factor 0 %, voxel dimension 2.0 x 2.0 x 2.0 mm³, TR = 7900 ms, TE = 90 ms, BW = 1502 Hz/Pixel, 40 diffusion encoding gradients, three intermittent non-weighted b0 images (b = 0 s/mm²), high b-value b = 1000 s/mm², GRAPPA with factor 2).

All images were investigated to be free of motion or ghosting and high frequency and/or wrap-around artefacts at the time of image acquisition.

**Supplementary Results**

**Image analysis**

**Supplementary Table 1 Association between FA-Values and MoCA**

| Negative Cluster | Location | Slope | Intercept | p-Value | Volume in mm³ | MNI152-Coordinates | | |
| --- | --- | --- | --- | --- | --- | --- | --- | --- |
|  |  |  |  |  |  | X | Y | Z |
| N1 | Left superior longitudinal fasciculus | -.0022 | .4609 | < .001 | 1289 | -32 | -19 | 21 |
| N2 | Right superior frontal gyrus  Middle frontal gyrus | -.0036 | .0976 | .021 | 231 | 25 | -1 | 54 |
| N3 | Left superior longitudinal fasciculus | -.0028 | .5169 | .033 | 218 | -39 | -43 | 11 |

**Supplementary Table 1.** Characteristics of clusters with an association between PD patients’ FA-values and postoperative change in MoCA. “Negative Cluster” denotes clusters with a negative association between patients’ FA-values and percentage difference of MoCA, i.e. higher FA-values were associated with lower postoperative change. “Location” indicates the anatomical landmark comprising the majority of voxels of a cluster according to Johns Hopkins University (JHU) white matter atlas, Harvard-Oxford cortical and subcortical atlas, and University College London (UCL) cerebellar atlas. P-Values are clusterwise p-values corrected for multiple comparisons. “Volume in mm³” denotes the size of a cluster and “MNI152-coordinates” describes the coordinates of the cluster’s centre of gravity in MNI152-space.

**Supplementary Table 2 Association between ODI-Values and MoCA**

| Positive  Cluster | Location | Slope | Intercept | p-Value | Volume in mm³ | MNI152-Coordinates | | |
| --- | --- | --- | --- | --- | --- | --- | --- | --- |
|  |  |  |  |  |  | X | Y | Z |
| P1 | Left superior longitudinal fasciculus  Corticospinal tract | .0016 | .2151 | < .001 | 469 | -32 | -13 | 29 |
| P2 | Right corticospinal tract Anterior thalamic radiation | .0012 | .2051 | < .001 | 225 | 26 | -7 | 21 |
| P3 | Right lingual Gyrus  Cerebellar V  Cerebellar VI | .0031 | .3259 | .011 | 153 | 19 | -51 | -14 |
| P4 | Right corticospinal tract | .0011 | .1336 | .029 | 136 | 25 | -15 | 24 |

**Supplementary Table 2.** Characteristics of clusters with an association between PD patients’ ODI-values and postoperative change in MoCA. “Positive Cluster” denotes clusters with a positive association between patients’ ODI-values and percentage differences in MoCA, i.e. higher ODI-values were associated with higher postoperative change. “Location” indicates the anatomical landmark comprising the majority of voxels of a cluster according to Johns Hopkins University (JHU) white matter atlas, Harvard-Oxford cortical and subcortical atlas, and University College London (UCL) cerebellar atlas. P-Values are clusterwise p-values corrected for multiple comparisons. “Volume in mm³” denotes the size of a cluster and “MNI152-coordinates” describes the coordinates of the cluster’s centre of gravity in MNI152-space.

**Supplementary Table 3 Association between NDI-Values and MoCA**

| Positive  Cluster | Location | Slope | Intercept | p-Value | Volume in mm³ | MNI152-Coordinates | | |
| --- | --- | --- | --- | --- | --- | --- | --- | --- |
|  |  |  |  |  |  | X | Y | Z |
| P1 | Left precentral gyrus | .0043 | .5943 | < .001 | 160 | -25 | -15 | 42 |
| P2 | Right Superior thalamic radiation | .0041 | .5896 | .003 | 136 | 19 | -5 | 39 |
| P3 | Right middle temporal gyrus, posterior division | .0062 | .3556 | .047 | 99 | 59 | -34 | -1 |
| Negative Cluster |  |  |  |  |  |  |  |  |
| N1 | Left lateral occipital cortex, inferior division  Inferior longitudinal fasciculus | -.0057 | .4382 | < .001 | 220 | -41 | -61 | 8 |
| N2 | Right lateral occipital cortex, superior division  Inferior fronto-occipital fasciculus  Inferior longitudinal fasciculus | -.0023 | .4949 | < .001 | 187 | 28 | -73 | 18 |
| N3 | Left lateral occipital cortex, superior division  Inferior longitudinal fasciculus | -.0049 | .4676 | .001 | 147 | -36 | -66 | 19 |
| N4 | Left cingulate gyrus, posterior division  Precentral gyrus | -.0049 | .2682 | .001 | 146 | -14 | -29 | 39 |
| N5 | Right superior longitudinal fasciculus | -.0043 | .6191 | .002 | 144 | 25 | -5 | 30 |
| N6 | Left precuneous cortex  Cingulum (cingulate gyrus) | -.0038 | .4855 | .014 | 114 | -16 | -62 | 34 |
| N7 | Right precuneous cortex  Cingulate gyrus, posterior division  Cingulum (cingulate gyrus) | -.0075 | .3420 | .026 | 106 | 7 | -50 | 34 |

**Supplementary Table 3.** Characteristics of clusters with an association between PD patients’ NDI-values and postoperative change in MoCA. “Positive Cluster” denotes clusters with a positive association between patients’ NDI-values and percentage differences in MoCA, i.e. higher NDI-values were associated with higher postoperative change. “Negative Cluster” denotes clusters with a negative association between patients’ NDI-values and percentage difference of MoCA, i.e. higher NDI-values were associated with lower postoperative change. “Location” indicates the anatomical landmark comprising the majority of voxels of a cluster according to Johns Hopkins University (JHU) white matter atlas, Harvard-Oxford cortical and subcortical atlas, and University College London (UCL) cerebellar atlas. P-Values are clusterwise p-values corrected for multiple comparisons. “Volume in mm³” denotes the size of a cluster and “MNI152-coordinates” describes the coordinates of the cluster’s centre of gravity in MNI152-space.

**Supplementary Figures**

**Supplementary Figure 1**

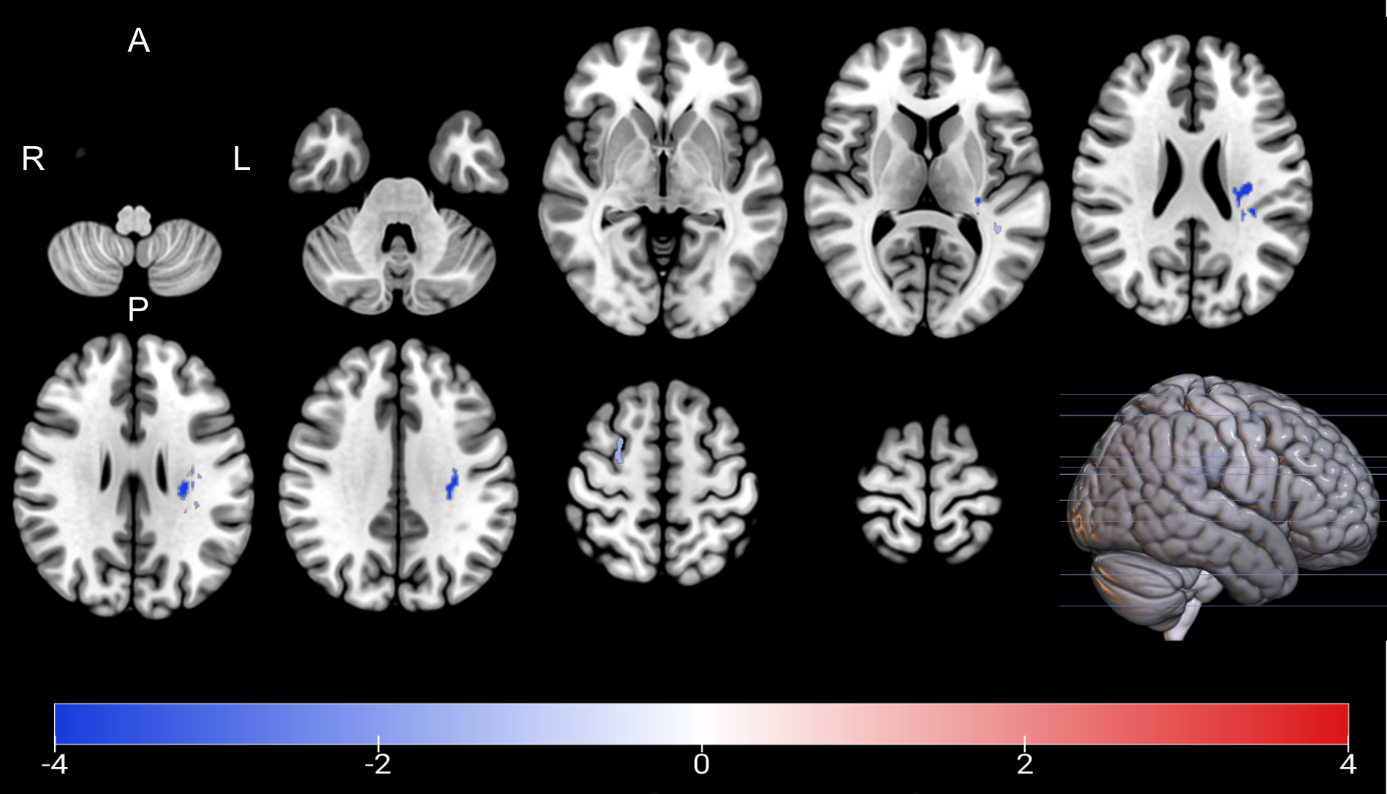

**Supplementary Figure 1. Association between FA and postoperative change in MoCA.** Clusters with a negative association between PD patients’ FA-values and postoperative change in MoCA as revealed by the whole brain analysis. Clusterwise p-values were corrected for multiple comparisons using a permutation-based approach and shown as the negative decadic logarithm of the p-value.

**Supplementary Figure 2**

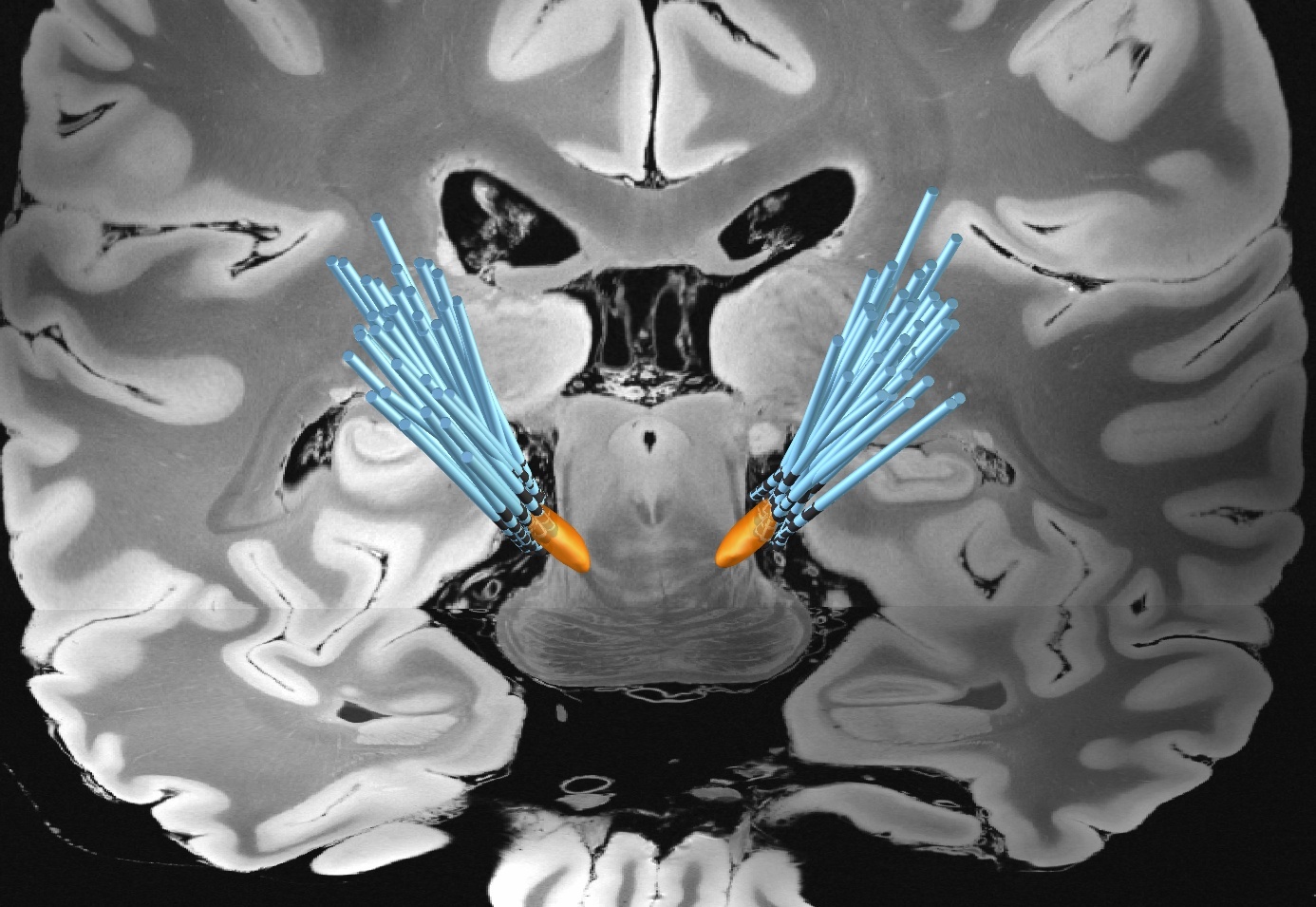

**Supplementary Figure 2.** reconstruction of the electrodes for the 40 patients as calculated within the Lead-DBS toolbox.

**Supplementary Figure 3**

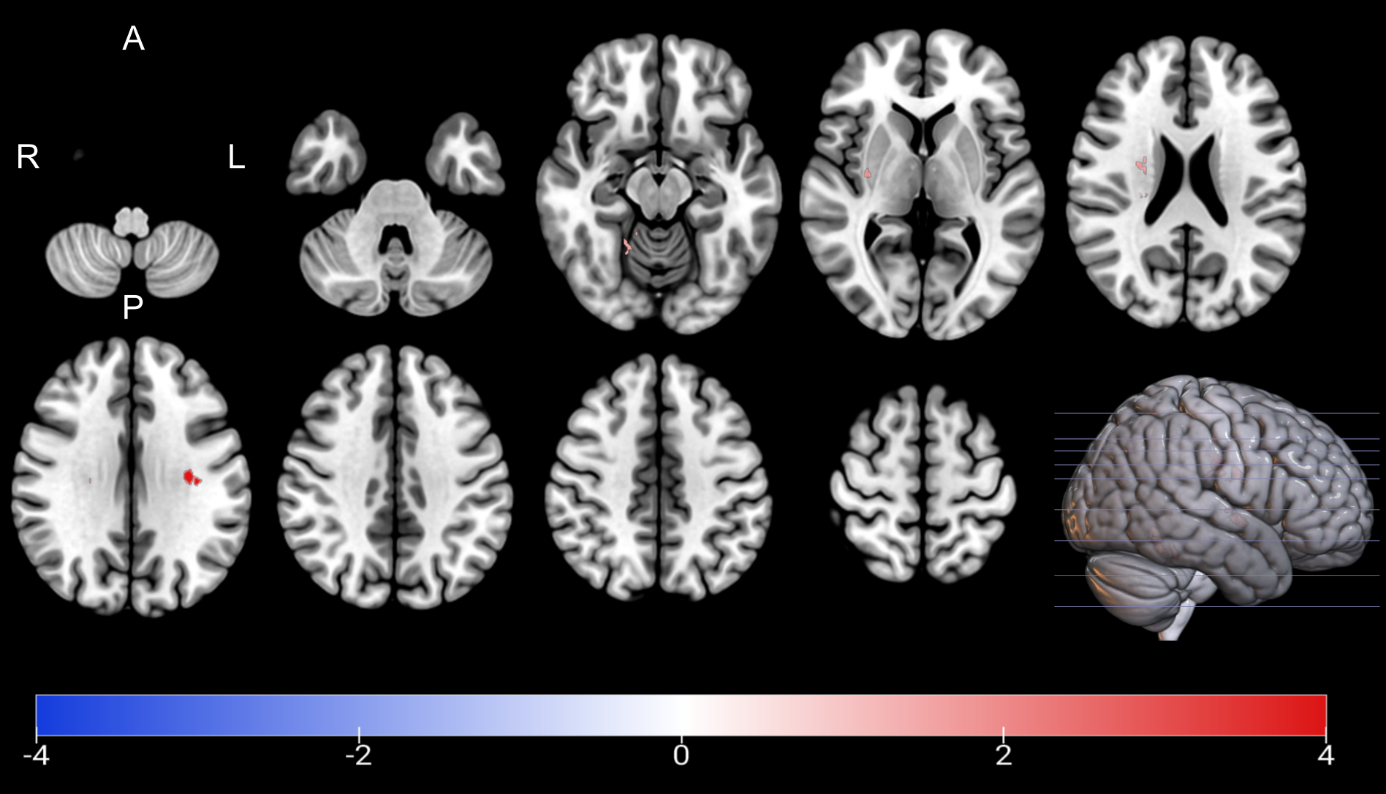

**Supplementary Figure 3. Association between ODI and postoperative change in MoCA with overlap between VTA and STN as covariate.** Clusters with a positive association between PD patients’ ODI-values and postoperative change in MoCA with overlap between VTA and STN as covariate as revealed by the whole brain analysis. Clusterwise p-values were corrected for multiple comparisons using a permutation-based approach and shown as the negative decadic logarithm of the p-value.

**Supplementary Figure 4**

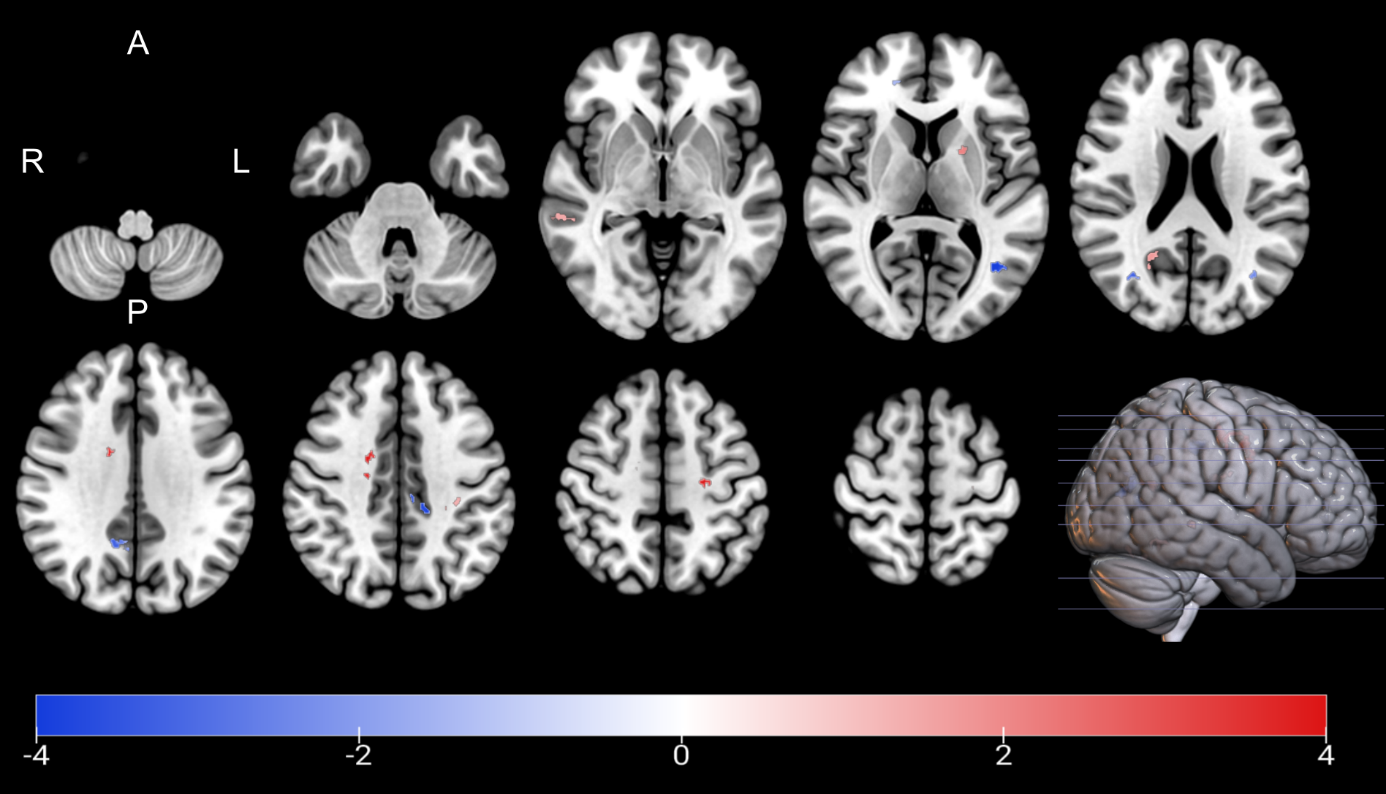

**Supplementary Figure 4. Association between NDI and postoperative change in MoCA with overlap between VTA and STN as covariate.** Clusters with a positive and negative association between PD patients’ NDI-values and postoperative change in MoCA with overlap between VTA and STN as covariate as revealed by the whole brain analysis. Clusterwise p-values were corrected for multiple comparisons using a permutation-based approach and shown as the negative decadic logarithm of the p-value.

**Supplementary Figure 5**

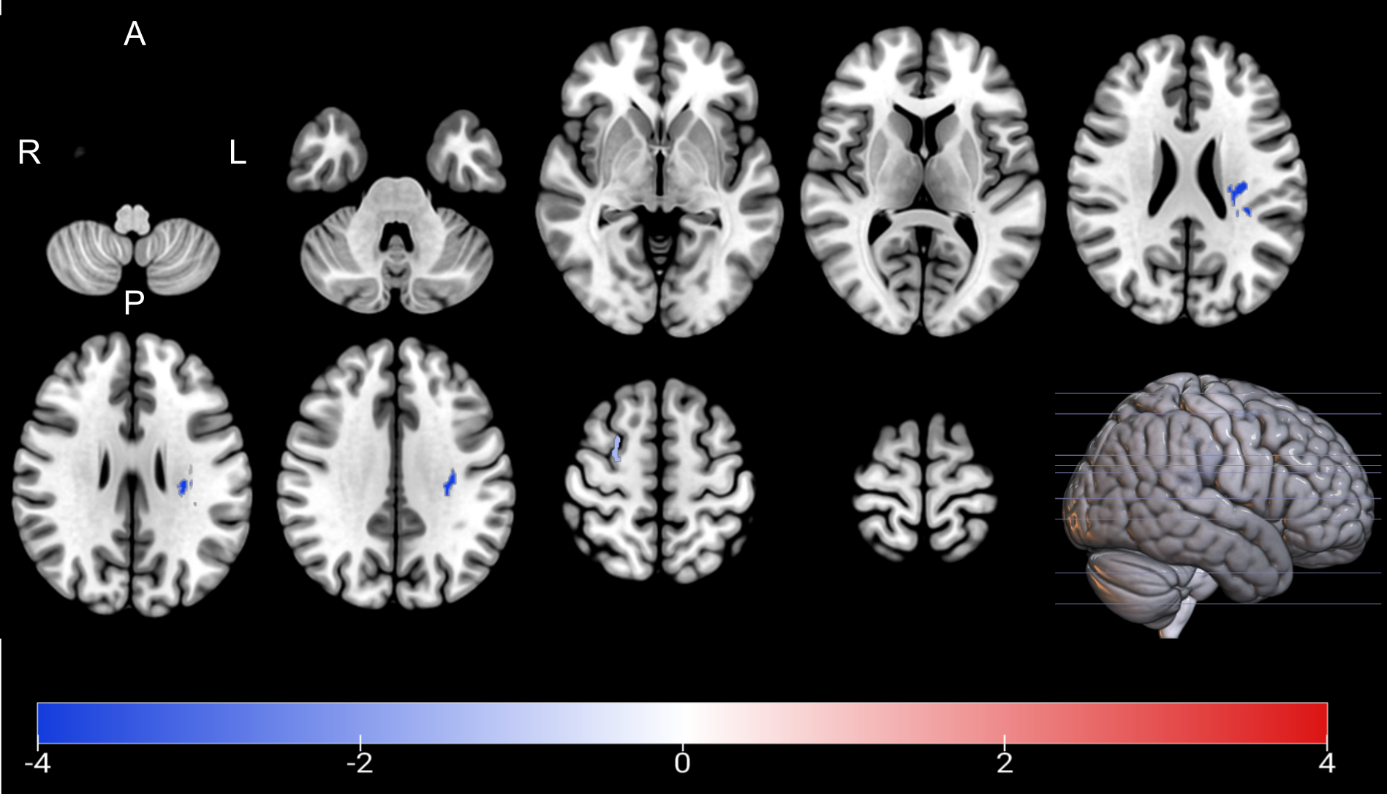

**Supplementary Figure 5. Association between FA and postoperative change in MoCA with overlap between VTA and STN as covariate.** Clusters with a negative association between PD patients’ FA-values and postoperative change in MoCA with overlap between VTA and STN as covariate as revealed by the whole brain analysis. Clusterwise p-values were corrected for multiple comparisons using a permutation-based approach and shown as the negative decadic logarithm of the p-value.
